# Patient ancestry significantly contributes to molecular heterogeneity of systemic lupus erythematosus

**DOI:** 10.1101/2020.05.31.20114660

**Authors:** Michelle D. Catalina, Prathyusha Bachali, Anthony E. Yeo, Nicholas S. Geraci, Michelle A. Petri, Amrie C. Grammer, Peter E. Lipsky

**Affiliations:** AMPEL BioSolutions LLC & RILITE Research Institute. 250 West Main Street, Suite 300. Charlottesville, Virginia; A. Yeo Consulting. Jersey City, New Jersey; Division of Rheumatology, School of Medicine, Johns Hopkins University, Baltimore, Maryland, USA

## Abstract

Gene expression signatures can stratify patients with heterogeneous diseases, such as Systemic Lupus Erythematosus (SLE), yet understanding the contributions of ancestral background to this heterogeneity is not well elucidated. We hypothesized that ancestry would significantly influence gene expression signatures and measured 34 gene modules in 1566 SLE patients of african (AA), european (EA) or native american (NAA) ancestry to determine the impact of ancestry on gene expression. Healthy subject ancestry-specific gene expression provided the transcriptomic background upon which the SLE patient signatures were built. Although standard therapy affected every gene signature, and significantly increased myeloid cell signatures, logistic regression analysis determined that ancestral background significantly changed 23/34 gene signatures. Additionally, the strongest association to gene expression changes was autoantibodies and this also had etiology in ancestry; the AA predisposition to have both RNP and dsDNA autoantibodies compared to EA predisposition to have only anti-dsDNA. A machine learning approach was used to determine a gene signature characteristic to distinguish AA SLE and was most influenced by genes characteristic of the perturbed B cell axis in AA SLE patients.

## Introduction

Systemic Lupus Erythematosus (SLE) is a complex, multigenic autoimmune disease affecting mostly women and characterized by autoantibodies to nucleic acids and nuclear proteins leading to immune complex formation, complement deposition and immune-mediated damage in multiple organ systems(1). The heterogeneity in ancestral prevalence, disease severity, organ involvement and response to treatment has been described, but the explanation has not been fully delineated(2). Therefore, the development of transcriptomic signatures to determine the basis of ancestral differences in lupus disease expression is of great interest. Whereas the disease is most prevalent in Asians and people of African-Ancestry(3–5) (AA), a disproportionate number of clinical trials have focused on the European Ancestry (EA) population(2, 6). Although not as extensively studied, native people of North American ancestry have also been shown to have earlier onset of disease and more organ involvement(7, 8). The Lupus in Minority populations: Nature vs Nurture (LUMINA) study and others have demonstrated increased active disease, organ involvement, and autoantibody levels for AA compared to EA patients(9, 10) and other studies have shown increased mortality for AA patients(11, 12). At the cellular level, the AA population has been shown to have more activated B cells, CD27-IgD-B cells, and B cell receptor signaling than the EA population(13). Several studies have demonstrated differences in responses of both innate immune cells as well as lymphocytes suggesting that ancestral differences in immune cells may contribute to the different disease course and incidence between populations(14, 15). Ancestry-related differences in response to therapy have also been reported. AA SLE patients responded better to B cell depletion therapies than Caucasian patients(16), but they displayed lesser responses to anti-BAFF treatment in a Phase III clinical trial(17, 18). Higher serum levels of BAFF in AA SLE patients have led to the suggestion that higher doses of the biologic may be necessary in AA patients(19).

Heterogeneity in SLE gene expression signatures were first reported for the IFN-stimulated genes(20, 21) and an association of IFN signatures with autoantibodies has been reported(22–28). Kirou et al(22) previously showed a significant association with anti-RNP, -Sm, -SSA and dsDNA autoantibodies with an interferon gene signature (IGS) and that patients having multiple autoantibodies also were more likely to have an IGS. Further work to describe SLE patient gene expression differences has been carried out by creating modules of genes over-represented in 158 pediatric SLE patients. Increased plasmablast, cell-cycle and erythroblast modules were detected in AA SLE patients and increased myeloid signatures and inflammation were observed in EA and Hispanic SLE patients suggesting that there may be an ancestral basis to explain some of the heterogeneity in SLE gene expression signatures(27). It is unknown whether adult SLE patients will have the same associations and whether other prominent gene expression signatures used to divide SLE patients into groups such as low density granulocytes, granulocytes, T cells, B cells, and platelets will also have gene expression differences based on ancestry(29).

Whole blood (WB) gene expression analysis provides a relatively straightforward means of assessing a subject’s transcriptomic fingerprint. We sought to determine the contribution of ancestry, sex, SoC therapy, serology and clinical manifestations to the WB gene expression profile of 1566 adult SLE subjects. This work provides strong evidence that much of the gene expression signature measured between SLE patients and healthy controls (HC) is related to patient ancestry resulting in alterations in the proportions of hematopoietic cells, cellular processes and signaling pathways detected. Importantly, the ancestry-related variance in gene expression in healthy persons contributes to the differences observed in subjects with SLE.

## RESULTS

### There is significantly different ancestral gene expression in SLE patients

In order to compare the ancestral contribution to gene expression, we made use of two large phase 3 clinical trial gene expression datasets (Illuminate (ILL) 1 and 2; GSE8884) with a minimum disease severity requirements of SLEDAI ≥ 6 and positive ANA that were well matched for average, median and range of SLEDAI and percentage of patients with anti-dsDNA between AA, EA and NAA SLE patients (**Supplemental Table 1**). These ancestral groups were also well matched for SLE manifestations used to determine SLEDAI (**Supplemental Table 2**)(30–32). Bulk differential expression (DE) analysis of ILL1 determined there were thousands of differentially expressed genes (DEGs) between 798 SLE patients of African, European and Native American (NAA) ancestry, but no differentially expressed transcripts when each ancestry was randomized into two groups and compared to itself. These differences were reproduced in a second cohort of 768 patients (ILL2), and then confirmed in another unrelated dataset (GSE45291) of 244 low disease activity AA and EA SLE patients that were also matched for mean, median and range of SLEDAI and ANA titer (**Supplemental Tables 3,4; Supplemental Figure 1**). We sought to determine how individual patient signatures contributed to these stable, reproducible group differences between ancestries. We employed gene set variation analysis (GSVA) with gene expression data from 1566 female AA, EA or NAA SLE patients (GSE88884 Illuminate 1 (ILL1) and Illuminate 2 (ILL2))(31) to compare enrichment of 34 gene modules corresponding to lymphocytes, myeloid cells, and cellular processes (**Figure 1A; Supplemental Table 5**(33)). We have previously used GSVA modules representative of cellular types and processes to determine enrichment in SLE patients and mice(33, 34). GSVA is advantageous compared to gene set enrichment analysis because it does not require a priori designation of two groups on the basis of phenotype and is advantageous when disease samples are highly heterogeneous and there are low numbers of control samples(35). GSVA demonstrated that NAA had the highest percentage of patients with enrichment of low density granulocyte (LDG), granulocyte, IL-1 and inflammasome signatures followed by EA patients, and AA had the lowest. NAA also had significantly more patients with enrichment of monocyte cell surface and monocyte modules than AA patients, but, notably, signatures for myeloid secreted proteins, which included complement components, *TNF*, and *CXCL10*, were not different between the three ancestries. AA had significantly more patients with B cell, immunoglobulin (Ig), plasma cell and T-regulatory (T-reg) signatures compared to EA and NAA. NAA patients had significantly fewer patients with T cell associated signatures compared to both EA and AA, whereas EA had significantly fewer patients with decreased dendritic cell (DC) and plasmacytoid DC (pDC) signatures compared to controls. The percentage of AA patients with enrichment of the IGS was higher than EA. AA and NAA had significantly fewer patients with decreased erythrocyte and platelet GSVA scores compared to EA (**Figures 1B,C Supplemental Table 6**). GSVA scores for the 34 cell and process modules were also calculated for 14 AA, 93 EA, and 17 NAA male patients and male HC in SLE dataset GSE45291. The pattern of enrichment was similar to that observed for the 1566 females in Figure 1B with increased plasma cells, Ig and T-reg signatures in AA SLE patients and increased LDG and myeloid signatures in and EA SLE patients, although statistical significance between the groups was noted only for the LDG, granulocyte, T-reg, TCRA, TCRB, and platelet signatures (**Supplemental Figure 2A; Supplemental Table 7**).

**Figure 1.**
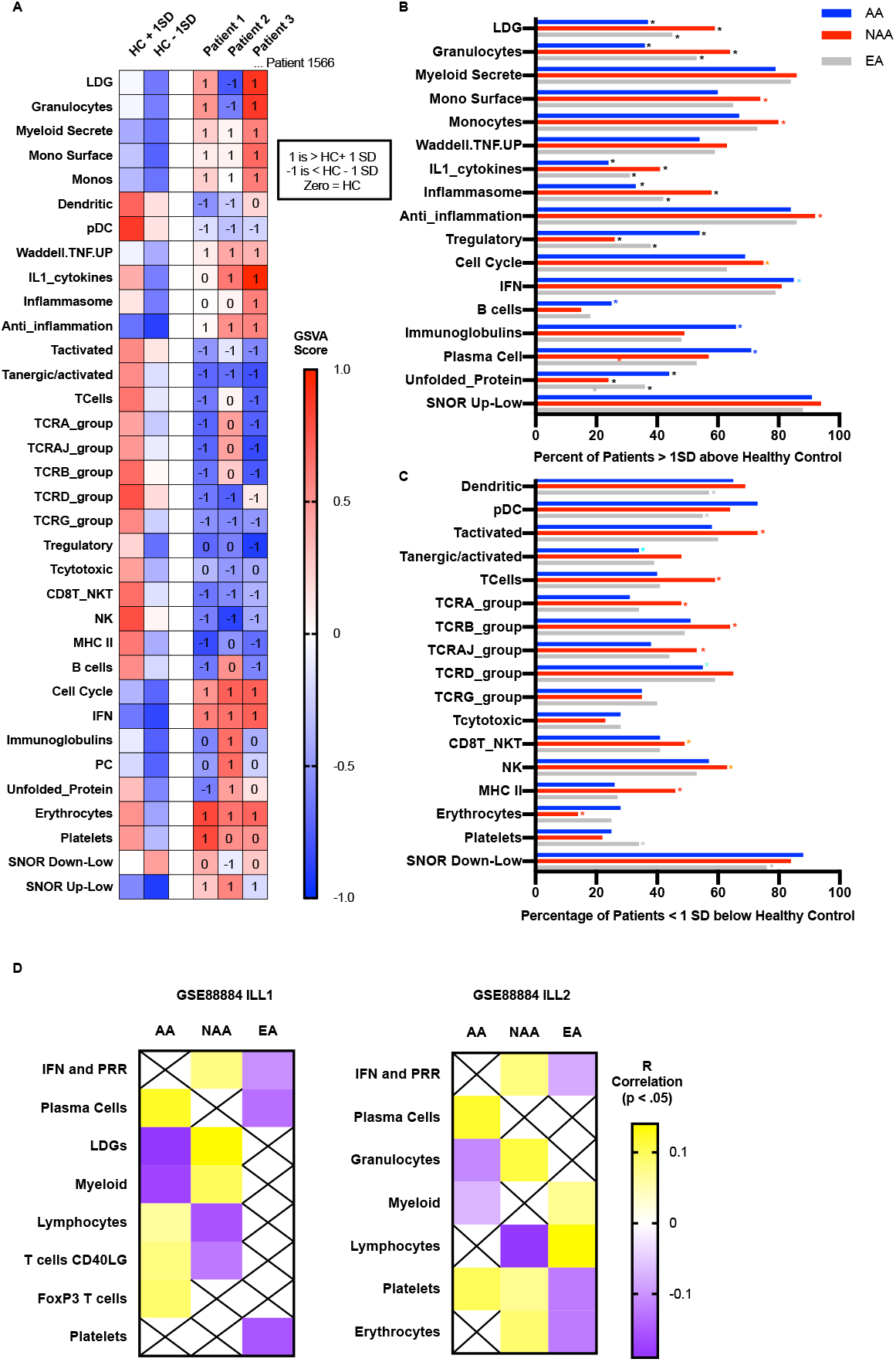
Ancestral bias is manifested by varied patterns of signatures for 34 cell and process modules. **(A)** Gene Set Variation Analysis (GSVA) was carried out on 17 female HC subjects to determine the mean and standard deviation (SD) of control GSVA scores for 34 cell type and process modules. HC mean scores plus or minus one standard deviation (SD) were used to determine a normal range for GSVA cell type scores. SLE female patient (GSE88884, ILL1 and ILL2 datasets; n = 1566) GSVA scores were determined and compared to HC values to determine whether patients had increased (+1), decreased (−1), or normal (zero) values. GSVA enrichment module gene symbols are in **Supplemental Table 5**. Percentage of patients within each ancestry (AA: n = 216, NAA: n = 232, EA: n = 1118) with **(B)** > or **(C)** < than 1 SD GSVA enrichment scores for each cell type and process module. Fisher’s exact p values < .05 are indicated by different color *: black * for comparisons to the two other ancestries, red * between NAA and AA/EA, orange * between NAA and EA, light blue * between AA and EA, dark blue * between AA and NAA/EA. Exact p values and percentages are listed in **Supplemental Table 6. (D)** WGCNA was carried out on dataset GSE88884 ILL1 and ILL2 cohorts separately. Pearson correlation r values to ancestry were determined for each module and listed if the p value was below 0.

Weighted gene co-expression network analysis (WGCNA) confirmed the association of ancestry with cellular signatures. WGCNA of female patients from the two cohorts of dataset GSE88884 was carried out separately and demonstrated a significant positive correlation of AA ancestry to plasma cell, T cell and T-reg gene modules and a significant negative correlation to granulocyte and myeloid cell modules. NAA ancestry exhibited positive correlations to IGS, granulocyte, platelet and erythrocyte modules and negative correlations to T cell and lymphocyte modules. EA ancestry was positively correlated to one myeloid cell module and negatively correlated to IGS, plasma cell, platelet and erythrocyte modules (**Figure 1D, Supplemental Table 8**). Thus, an orthogonal approach using co-expression defined gene clusters confirmed the ancestral-related gene expression differences.

### Ancestry provides the gene expression backbone for SLE gene expression abnormalities

Analyses of DEGs detected between different ancestries showed that AA populations had decreased expression of the Duffy blood group antigen *ACKR1*, the platelet, dendritic, and monocyte receptor *CD36*, and *G6PD* in comparison to NAA and EA populations (**Supplemental Table 4**) and these genes have previously been described as risk alleles resulting in decreased expression in AA(36–38). We hypothesized that ancestral-related gene expression differences detected between SLE patients may be related to heritable differences in expressed genes in hematopoietic cells of healthy subjects. In order to address this question, DE analysis was carried out between AA and EA healthy subjects from two separate datasets (**Supplemental Table 9**) and compared to the DEGs that differed between AA to EA SLE patients. There was a highly significant overlap in transcripts differentially expressed between healthy AA and EA subjects and transcripts differentially expressed between AA and EA SLE patients (**Figure 2A**). GSVA was carried out on the healthy AA and EA subjects and enrichment scores were compared for the 34 cell and process modules. Ten of the 34 signatures were significantly different between AA and EA healthy subjects. Healthy EA subjects had significantly increased granulocyte, inflammasome, monocyte cell surface, monocyte, inflammatory secreted and dendritic cell GSVA enrichment scores compared to AA healthy subjects, and AA healthy subjects demonstrated significantly increased T activated, B cell, erythrocyte and platelet GSVA enrichment scores compared to healthy EA subjects. No differences in LDG, plasma cell, T cell, IGS or the other signatures were determined (**Figure 2B**). Thus, in the absence of disease, significant and reproducible gene expression differences exist between AA and EA and appear to be contributing to the molecular heterogeneity in gene expression.

**Figure 2.**
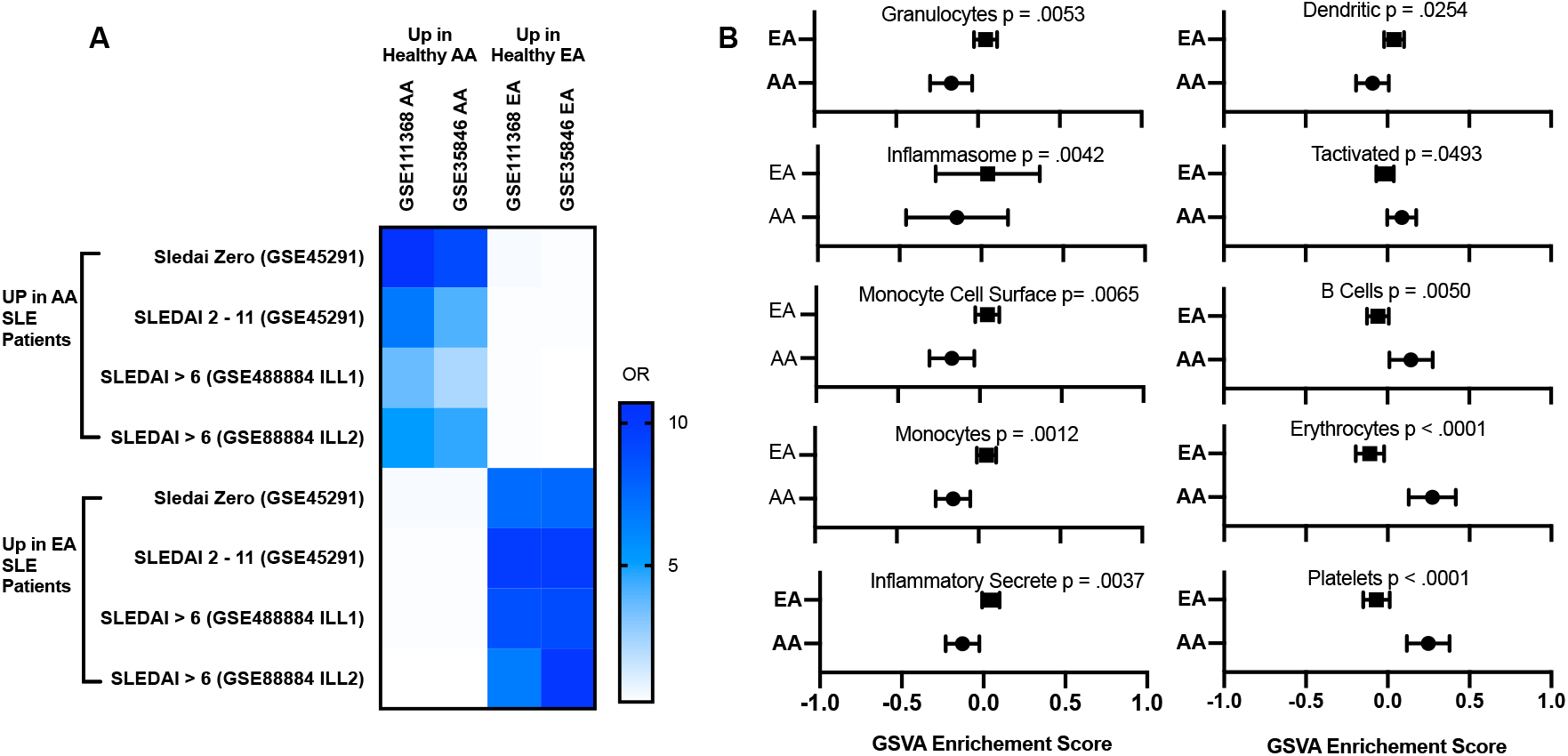
Gene expression differences in SLE patients are similar to ancestral gene expression differences in healthy controls. **(A)** Limma DE analysis was carried out between HC AA and EA for two separate datasets. Increased (Up in AA) and decreased (Up in EA) transcripts were compared to four SLE datasets of AA DE to EA. Overlap p values were all below 10E-22 for OR above 1. **(B)** GSVA for the 34 cell and process modules **(Supplemental Table 5**) was carried out on healthy AA and EA subjects from two separate datasets. Welch’s t-test was used to determine significant differences between ancestral GSVA scores and the mean and confidence intervals for the 10 GSVA scores significantly different (p < .05) between ancestries are shown.

### Autoantibodies and complement levels, but not other clinical features of lupus were associated with significant changes in gene expression profiles

Variation in SLE disease manifestations has been reported as a potential cause for gene expression heterogeneity in SLE WB(27, 28, 39). However, the presence of arthritis, rash, alopecia, mucosal ulcers or vasculitis had no consistent effect on cellular and process gene enrichment scores. Patients of all ancestries with both anti-dsDNA and low C had significantly higher GSVA scores for anti-inflammation, IGS, plasma cell, Ig, monocyte cell surface and LDGs compared to patients without anti-dsDNA and low C. (**Figure 3**).

**Figure 3.**
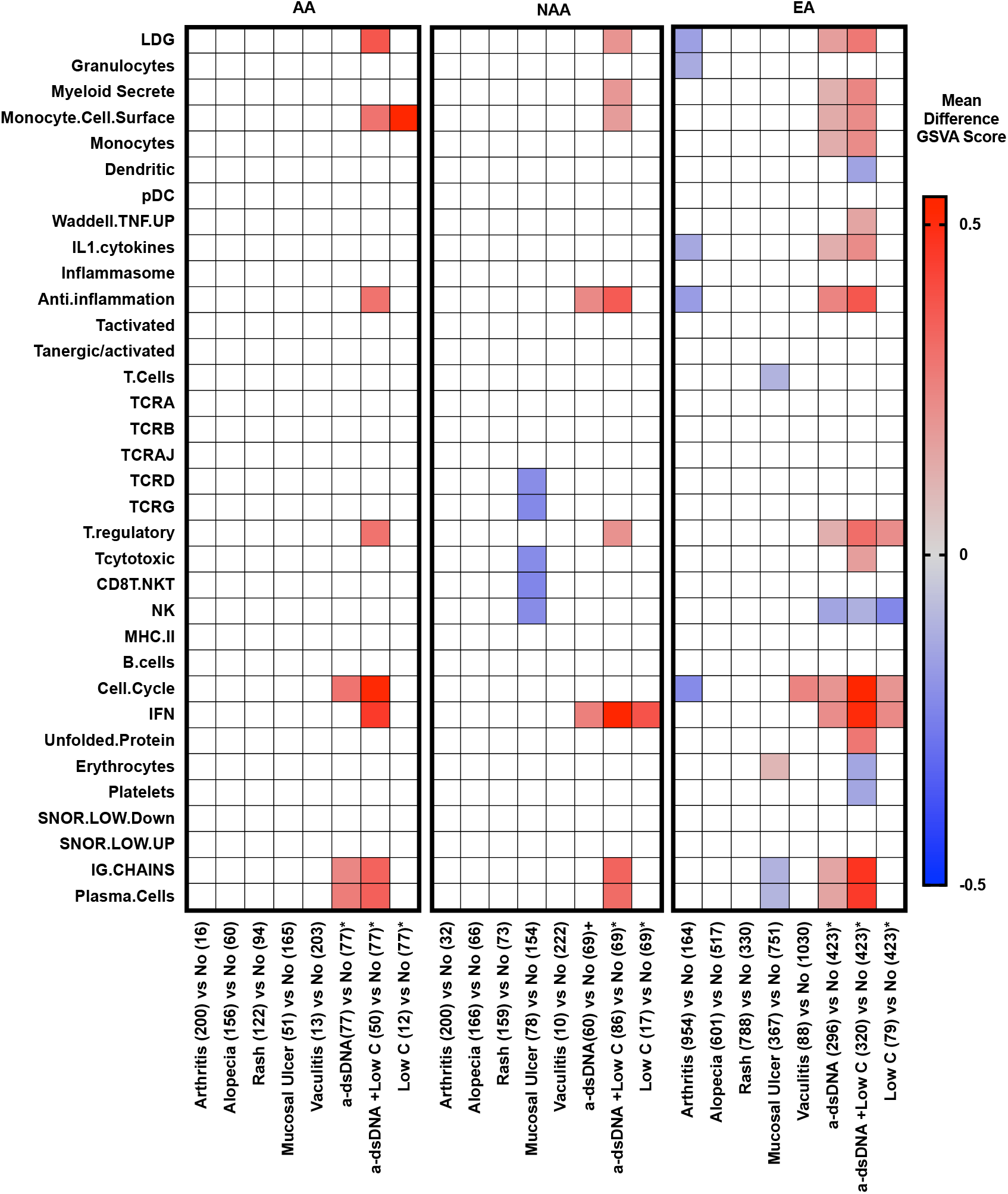
Autoantibodies and complement levels were associated with gene expression profiles. Number of patients with each SLEDAI component manifestation are shown in parentheses. Sedak’s multiple comparisons test was used to determine whether differences exist in gene expression signature GSVA scores for SLE patients with specific manifestations compared to all other manifestations. The mean difference in GSVA enrichment scores is shown for manifestations with significant (p < .05) differences in enrichment scores as compared to all other manifestations. *For anti-dsDNA autoantibodies and low C, patients were compared to patients without either anti-dsDNA (IU < 30) or low C (C3 > .8 g per L and C4 > 0.1 g per L). All patients in these analyses were positive for ANA.

Notably, the significant increase in plasma cell signatures detected in AA patients could not be explained by AA SLE patients having an increased incidence of anti-dsDNA and low C; AA had the lowest number and percentage of patients with both anti-dsDNA and low C (23%), whereas 29% of EA and 37% of NAA had anti-dsDNA and low C. Anti-RNP and anti-Sm autoantibodies have been demonstrated to be increased in SLE patients of African ancestry(13, 40–42) and these autoantibodies could also be related to plasma cell, IFN and other gene expression signatures. To understand how multiple autoantibodies change the transcriptome, we first determined the combinations of the five autoantibodies measured in this study for 1535 of the female SLE patients from ILL1 and ILL2: anti-dsDNA, anti-ribonucleoprotein (RNP), anti-Sm, anti-SSA, and anti-SSB. AA and NAA SLE patients had significantly higher frequencies of autoantibodies that are not dsDNA. Significantly fewer AA and NAA SLE patients were negative for all five autoantibodies compared to EA SLE patients. AA SLE patients had a significantly higher percentage with three or four autoantibodies and a significantly lower percentage of patients with only one autoantibody compared to EA, but there were no significant differences between AA and NAA. NAA SLE patients had significantly higher percentages of patients with four or five autoantibodies compared to EA (**Figure 4A, Supplemental Table 10**). Importantly, the presence of multiple autoantibodies was associated with significantly higher frequencies of the IGS and the plasma cell signature (**Figure 4B**).

**Figure 4.**
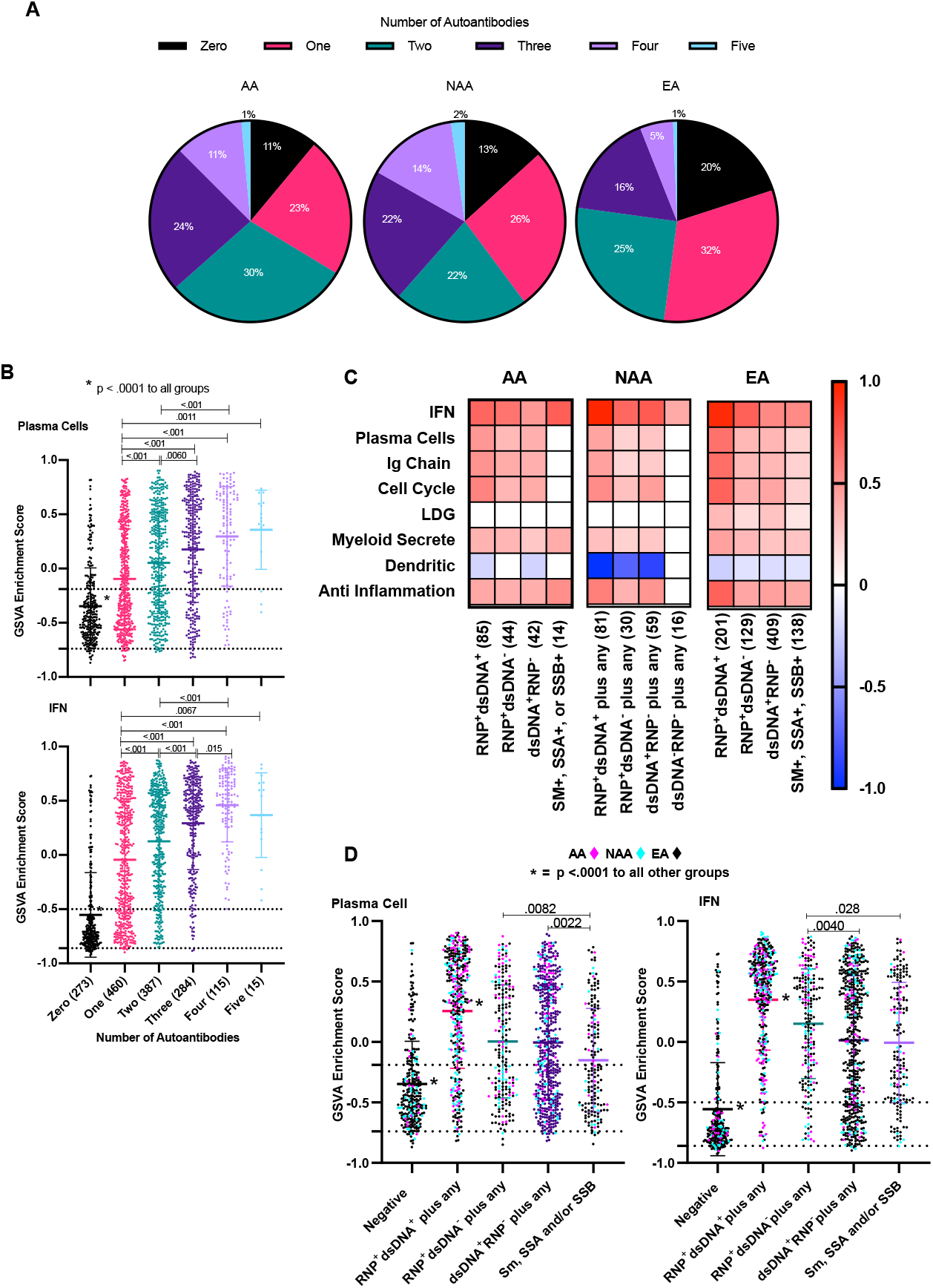
The higher number and different types of autoantibodies in AA SLE patients led to higher plasma cell, IGS, cell cycle, T-regulatory and myeloid secreted signatures. **(A)** Percentage of patients with different numbers of autoantibodies (RNP, Sm, SSA, SSB, and dsDNA) by ancestry. Borderline titers were considered negative for these analyses. **(B)** Comparison of plasma cell and IGS GSVA scores by patient number of autoantibodies. Patient n values are given in parentheses. Tukey’s multiple comparisons test was used to determine significant differences between GSVA scores for plasma cells and IGS for each group; p values < .05 are shown on the graph. **(C)** GSVA enrichment scores for all 34 cell and process modules were compared for each autoantibody group to patients of the same ancestry with 0/5 autoantibodies. Tukey’s multiple comparisons test was used to determine significant differences; eight cell and process module signatures with significant differences (p < .05) in GSVA scores are shown. **(D**) GSVA enrichment scores for plasma cells and IGS for AA, NAA and EA SLE patients combined into five autoantibody groups (1) 0/5 autoantibodies, (2) RNP+dsDNA+ plus any other, (3) RNP+dsDNA-plus any other, (4) RNP-dsDNA+ plus any other, (5) RNP-dsDNA-plus Sm, SSA or SSB. **(B,D)** Dots represent single patient scores and the error bars are mean and standard deviation. Numbers of patients in each group of autoantibodies are shown in parentheses. The black dotted lines represent the mean plus or minus 1 SD of the HC for GSVA scores.

For all three ancestries, patients positive for both anti-RNP and anti-dsDNA plus any of the other three autoantibodies had significantly increased enrichment scores for plasma cells, IGS, Ig, cell cycle, T-reg, myeloid secreted and anti-inflammation signatures compared to SLE patients negative for all five autoantibodies. (**Figure 4C**). Additionally, patients positive for anti-RNP plus any of the other autoantibodies except anti-dsDNA had significantly increased plasma cell and IGS GSVA scores compared to patients positive for anti-dsDNA plus any other autoantibody, and patients with any combination of anti-Sm, SSA and SSB (**Figure 4D**). This data explained the significantly increased plasma cell and IFN enrichment scores for AA SLE patients. AA SLE patients had significantly higher percentages of patients with anti-RNP autoantibodies (62%) compared to EA (30%) and NAA (51%), and significantly higher percentages of patients with anti-Sm (24%) compared to EA (12%), (**Supplemental Table 11)**. AA also had significantly increased numbers of patients with both anti-RNP and anti-dsDNA compared to EA, and significantly increased numbers of patients with anti-RNP+anti-dsDNA-plus anti-Sm, SSA or SSB autoantibodies compared to EA and NAA. AA and NAA also exhibited more frequent SM, SSA, or SSB autoantibodies compared to EA. (**Supplemental Table 12**). This data confirms in a large cohort of AA, EA and NAA SLE patients ancestrally-related disparities in autoantibody profiles and extends those findings to indicate that there is a significant association between autoantibody profiles and differences in gene expression between ancestries.

Autoantibody patterns in male SLE patients were similar to those determined in females, although statistical significance was not determined because of low patient numbers (**Supplemental Table 13**). Similar to female SLE patients, significantly increased IGS GSVA scores were determined for males with anti-RNP and anti-dsDNA plus any of the other three autoantibodies, and with RNP+dsDNA-versus anti-dsDNA+ plus any of the other three autoantibodies, and all of these groups were significantly different from patients with none of these five autoantibodies (**Supplemental Figure 2B**).

### Standard of Care (SoC) therapy is associated with significant changes in gene expression profiles

SoC therapy has been demonstrated to significantly affect SLE gene expression signatures(27, 43) and significantly more NAA SLE patients were receiving corticosteroids (92%) and taking immunosuppressives (IS) (58%) compared to 70% and 39% of AA and 70% and 39% of EA patients, respectively (Fisher’s exact p < .0001). It was, therefore, important to consider therapy affects on gene expression and determine whether ancestry or SoC drugs or both were contributing the differences in gene expression profiles. Corticosteroids significantly increased LDG and anti-inflammation GSVA scores compared to patients of the same ancestry not taking the drugs. Additionally, both AA and EA receiving corticosteroids had significant enrichment for granulocytes, myeloid secreted, monocyte cell surface, monocytes, cell cycle and the IGS. The effect of corticosteroids on myeloid signatures was further amplified at corticosteroid doses > 15 mg/day. When IS therapy was restricted to just MMF and MTX, there was a consistent decrease across all three ancestries in plasma cell and Ig GSVA scores. AZA significantly decreased NK cell GSVA scores in all three ancestries and also significantly decreased T cytotoxic and B cell scores in NAA and EA ancestries. EA patients receiving NSAIDs compared to all other treatments had decreased LDG and anti-inflammation signatures, whereas anti-malarials had no significant effect on GSVA enrichment scores (**Figure 5**). Two separate cohorts of SLE patients with low disease activity from dataset GSE45921 also had SoC drug information and were analyzed to confirm the findings. Corticosteroids increased LDG, monocyte and anti-inflammation GSVA scores; MTX and MMF decreased plasma cell GSVA scores; and AZA decreased NK and B cell GSVA scores (**Supplemental Figure 3**) in support of the data generated with the first dataset composed of 1566 female SLE patients.

**Figure 5.**
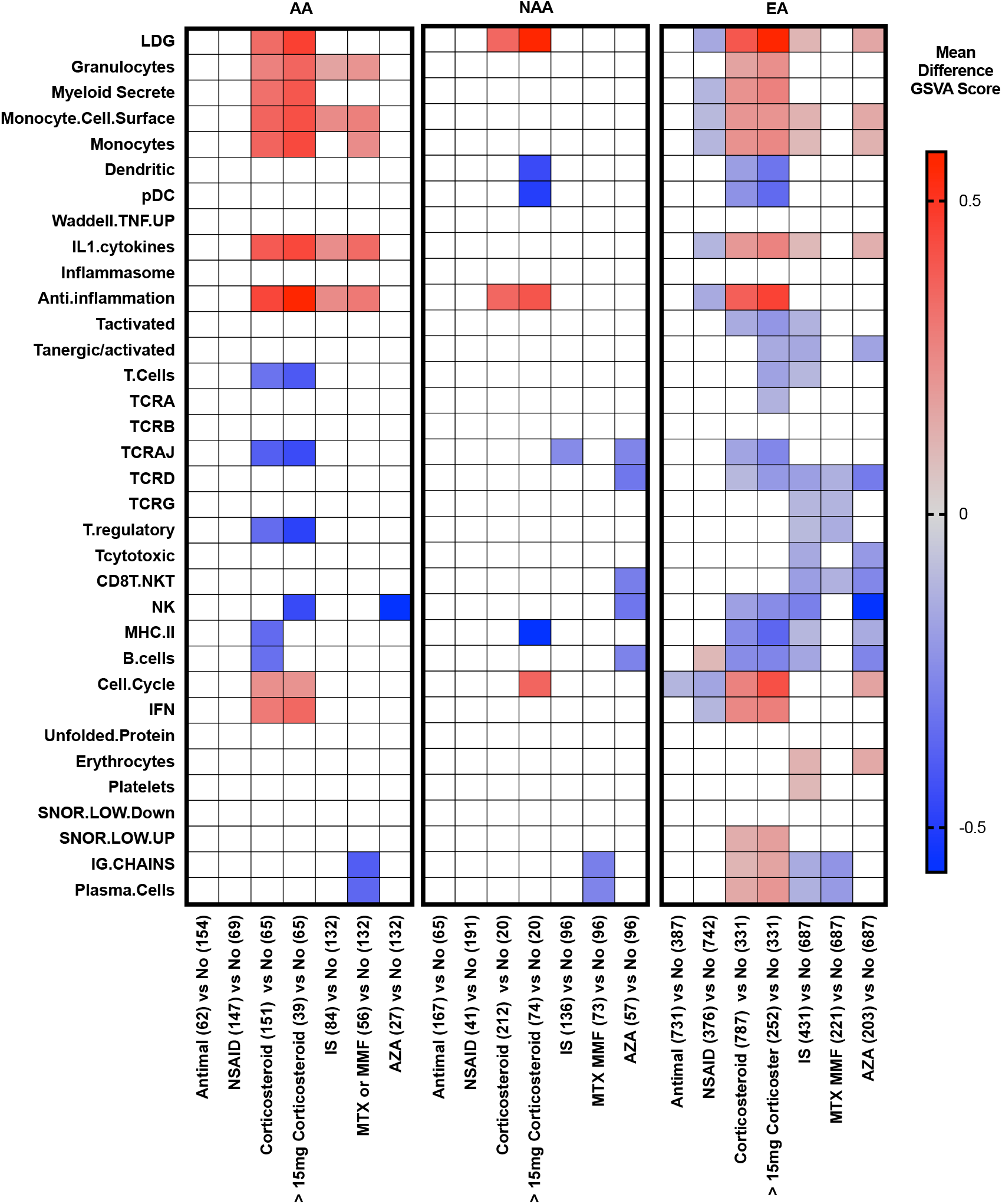
Association of corticosteroid use and immunosuppressive therapy with changes in gene expression profiles. 1566 female SLE patients (GSE88884) were separated by ancestry and GSVA scores for each cell type or process module in patients receiving each therapy were compared to GSVA scores for each cell type or process module in patients taking all other therapies. The patient numbers are in parentheses. Sidek’s multiple comparisons test was used to determine significant differences for the mean difference in GSVA scores between therapies. The mean difference in GSVA score related to the treatment is shown for therapies with p values < .05. Two EA patients were receiving cyclophosphamide and are included in the immunosuppressive (IS) calculation for EA category.

### Sex has a less important effect than ancestry on gene expression differences

Because of the large number of EA females, we were able to balance the percentage of female and male patients on corticosteroids and IS in order to determine gene expression differences between male and female EA SLE patients (**Supplemental Table 15**). We also divided the females into two age groups, 25 – 49 years and > 50 years, because of the reported effects of estrogen on immune responses(44). There were very few differences between male and female SLE patients in gene expression (**Supplemental Figure 4, Supplemental Table 16**) suggesting that ancestral differences are a more important factor in gene expression than sex differences.

### Logistic regression modeling demonstrated that ancestry is the major influence on SLE gene expression differences

To determine the relative importance of ancestry, SLE manifestations, serology and SoC drugs on gene expression signatures, we performed stepwise logistic regression on data from 1535 female SLE patients with all five autoantibody measurements for each of the 34 cell type and process signatures using the variables of ancestry, SoC drugs, SLE serologic abnormalities, SLE manifestations, age, and time from onset of disease. Co-linearity was excluded by carrying out Spearman correlations between all variables and the ethnic term Hispanic was removed from modeling because of an r_s_ of 0.54 to NAA (**Supplemental Table 17**). Figure 6 shows CIRCOS visualizations of the odds ratios (OR) for each variable significantly contributing to each GSVA score. Ancestry was associated with changes in 23 cell or process signatures. AA ancestry was positively associated with T-reg, plasma cell, Ig, and low pDC signatures and negatively associated with granulocyte, monocyte, IL1, anti-inflammation, and low B cell signatures. NAA ancestry had the highest positive association to the inflammasome and a negative association to T-reg signatures. NAA was also positively associated with erythrocyte, low T cell and low MHC II, and negatively associated with the IGS and unfolded protein response signatures. EA was positively associated with high myeloid secreted, inflammasome, and low platelet, and negatively with low NK and T-reg signatures (**Figure 6A, Supplemental Table 18**). SLE serologic profiles are interrelated to ancestral background and had the highest OR to significant changes in GSVA scores. Autoantibody groups RNP+dsDNA+, RNP+dsDNA-, RNP-dsDNA+, and any combination of Sm, SSA and SSB resulted in significant OR of 31.6, 25.6, 5.5 and 13.1, respectively, for the relationship to the IGS; OR of 7.9, 4.7, 4.0, and 2.3 respectively for the relationship to the cell cycle signature; OR of 8.7, 3.9, 3.5, and 2.4 respectively to the plasma cell signature; OR of 4.8, 3.0, 2.4 and 2.2 respectively to the T-reg signature; OR of 3.6, 2.4, 2.4 and 2.1 to the TNF signature, and OR of 9.0, 3.5, 2.6 and 3.4 to the myeloid secrete signature. In total, autoantibodies and low C were related to changes in 23 cell and process signatures (**Figure 6B, Supplemental Table 19**). SoC drugs influenced every cell and process module GSVA score. Corticosteroids were significantly associated with increases in 14 cell and process signatures; the highest OR was 3.8 to the LDG signature. AZA was significantly associated with 9 signatures and had an OR of 4.8 to low NK cell signatures. Both MTX and MMF were associated with decreased lymphocyte signatures, especially plasma cells with OR of .394 and .211 respectively. (**Figure 6C, Supplemental Table 20**). Time, age and clinical manifestations were associated with the fewest changes and the lowest ORs. Age > 50 was related to changes in 12, and the time from onset of disease (TMONSET) was related to changes in nine of the 34 cell type and process modules. Notably, a time of onset of < one year was negatively associated with low B cells and age greater than 50 was negatively associated with the plasma cell signature. Clinical manifestations were related to changes in 17 cell type and process modules with mucosal ulcers related to changes in 13 modules; predominantly those associated with low T cell signatures. (**Figure 6D, Supplemental Table 21**).

**Figure 6.**
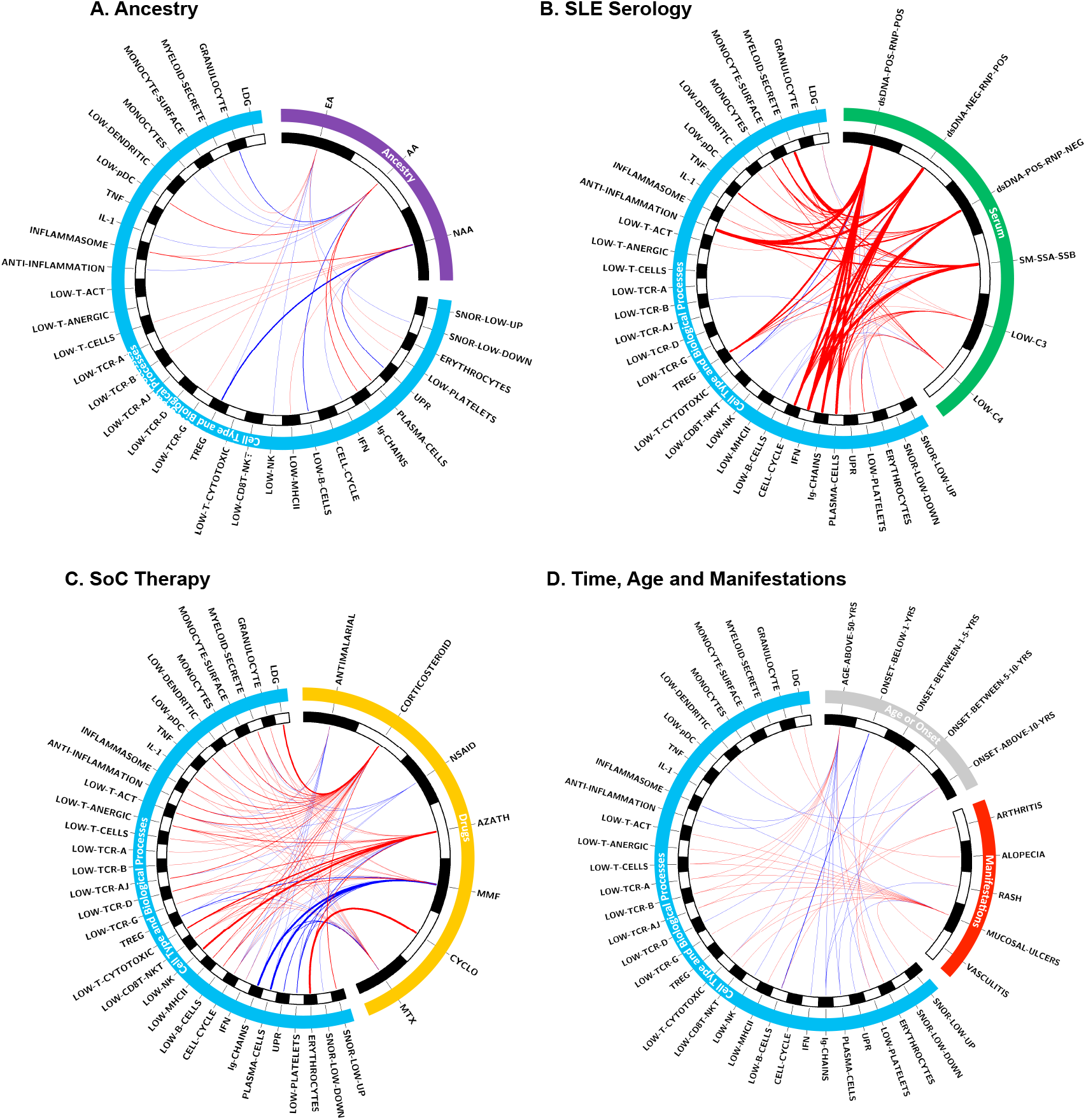
Stepwise logistic regression analysis determined the importance of ancestry, SoC drugs and SLEDAI components to the whole blood gene expression profile. Circos visualization of OR generated using stepwise logistic regression and showing the contributions of 26 variables to each of the 34 GSVA enrichment scores for cell type and process modules. The thickness of the lines connecting the 26 variables to the GSVA scores represent the magnitude of the odds ratios and an interval graph was used to assign thickness of the lines where OR < 2 = 1pt; 2 ≥ OR < 3 = 5pt; 3 ≥ OR < 10 = 10pt; OR ≥ 10 = 20pt. Red lines indicate odds ratios above 1 and blue lines indicate odds ratios below 1. OR between 0 and 1 are represented as 1/odds ratio to accurately reflect the magnitude of the negative relationship to the GSVA enrichment score. **(A)** Ancestral relationships to cell and process modules GSVA scores with p values < .05 (p values, OR, and CI in **Supplemental Table 18). (B)** Serology (autoantibody and complement) relationships to cell and process module GSVA scores with p values < .05 (p values, OR, and CI in **Supplemental Table 19). (C)** SoC drug relationships to cell and process module GSVA scores with values < .05 (p values, OR, and CI in **Supplemental Table 20). (D)** Time from onset of disease (TMONSET), age > 50, and SLE manifestation relationships to cell and process module GSVA scores with p values < .05 (p values, OR, and Cl in **Supplemental Table 21)**.

### Machine Learning Identifies the Perturbed B Cell Axis in AA SLE

Ancestry was associated with significant changes in 23 of 34 gene expression modules and, additionally, the high OR for association of gene expression signatures with serologic components suggested that one aspect of ancestry was to bias the tendency to form multiple autoantibodies, including anti-RNPs. Comparison of GSVA enrichment scores of patients with and without specific therapies confirmed the logistic regression results, indicating that while therapy had an important influence, ancestry was still a major contributor to gene expression profiles. To confirm this conclusion we carried out a machine learning approach to determine whether gene expression could predict African ancestry in SLE and also to determine the major predictors of ancestry. Because NAA signatures in this study were biased by substantial drug therapy, they were not used, whereas AA and EA had similar drug therapy profiles (**Supplemental Table 22**)

Logistic regression and two different machine learning algorithms were used to distinguish AA SLE patients from EA SLE patients using the gene expression values for the list of 752 genes comprising the modules used for GSVA (**Supplemental Table 5**). Logistic regression analysis, an elastic generalized linear model (GLM), and Support Vector Machine (SVM) were deployed to predict the ancestry status of SLE samples and determine the top 25 predictors using the gene importance score. All three models showed good performance with minor differences in their highest and lowest accuracies in each dataset. The SVM classifier was the strongest performer with 97 and 96 percent accuracy in Illuminate1 and Illuminate2, respectively. To ensure that models were not picking the noise, while learning the details in the training data, 10-fold cross validation was performed on each dataset separately and also combining the two datasets together. In both cases, the SVM outperformed the other classifiers with accuracy of 96 percent accuracy (**Figure 7A, Table 1**). The genes used to classify AA SLE compared to EA SLE reflect the perturbed B cells axis in AA SLE (**Figure 7B**). In a separate analysis, the same approach was used with the entire Illuminate data sets including the NAA subjects and very similar results were obtained (**Supplemental Figure 6**).

**Table 1.**
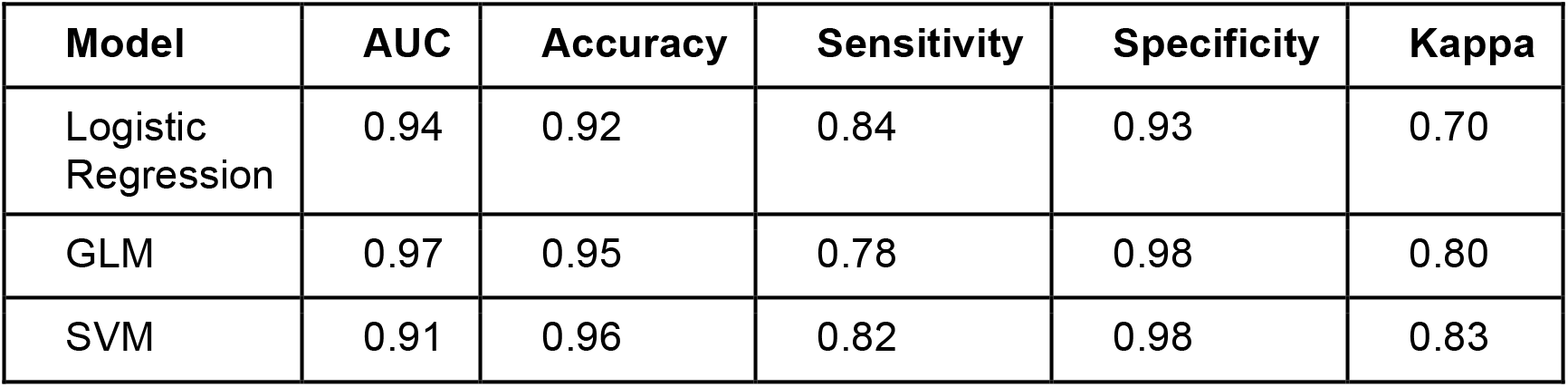
Classification metrics of machine learning classifiers.

**Figure 7.**
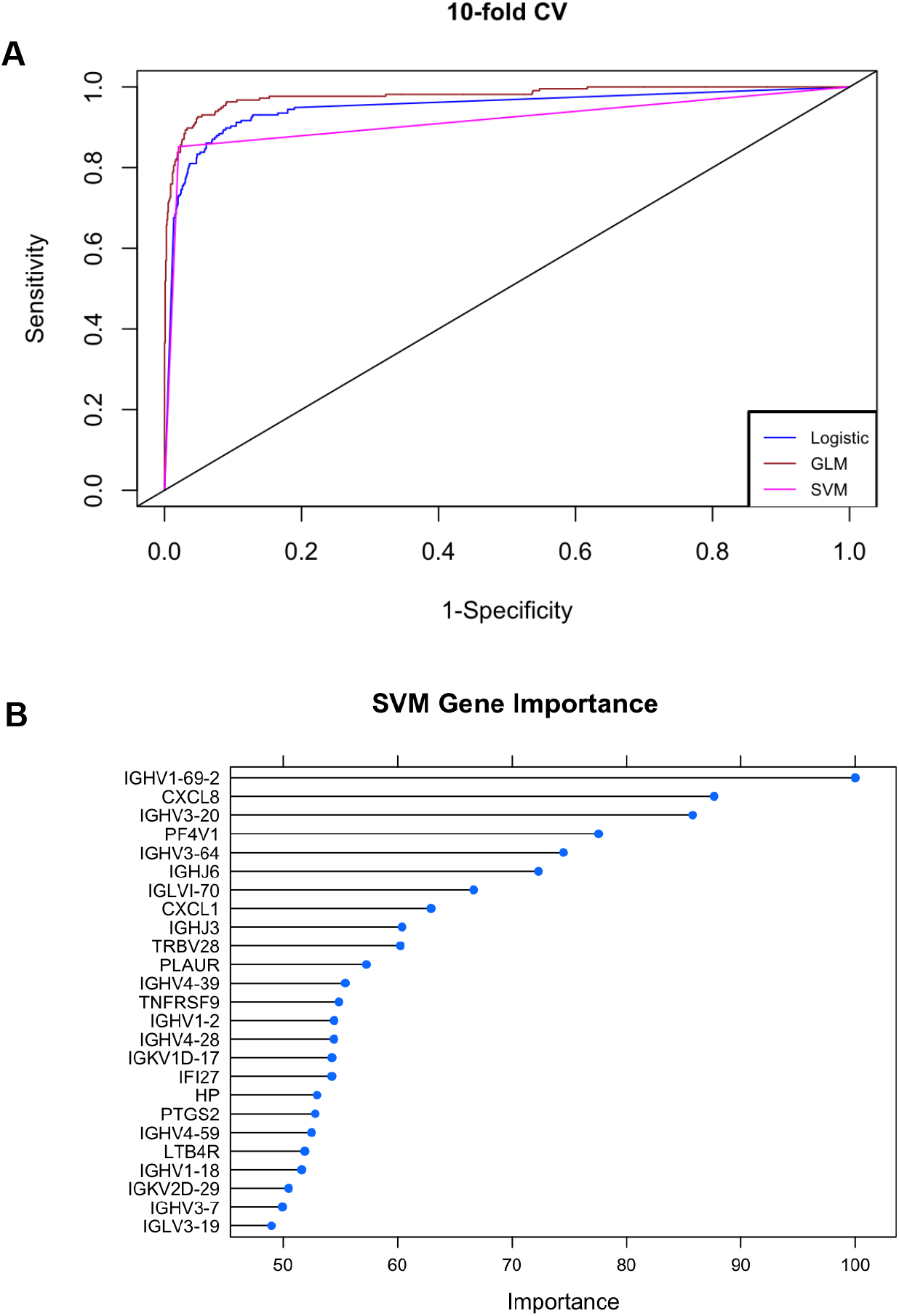
A machine learning approach predicted AA from EA SLE patients and demonstrated the perturbed B cell axis in AA SLE. **(A)** SLE patients were classified as African American (AA) using logistic regression, generalized linear models (GLM), and support vector machine (SVM) classifiers. ROC curve for logistic regression and the two different machine learning models in GSE88884 (ILL1 and ILL2 combined). **(B)** Top 25 predictors determined by SVM model.

## Discussion

This work demonstrated the significant impact of ancestry on gene expression patterns in SLE and by implication on the biologic pathways driving disease in patients of each ancestry. The increased plasma cell, IFN, Treg and inflammatory cytokine signatures were most strongly related to the AA ancestral bias of having increased anti-RNP/SM autoantibodies and multiple autoantibodies. Additionally, AA was independently associated with plasma cells and Ig transcripts when modeled alongside autoantibodies suggesting that AA SLE patients may have higher background levels of plasma cells. Furthermore, machine learning algorithms accurately identified AA SLE patients from their gene expression data and identified genes associated with B cells as important for distinguishing AA SLE. This is further evidence of the perturbed B cell lineage described in AA SLE patients(13, 19, 25, 45), and related to the increase in the B cell axis that was detected in healthy AA and might translate into a greater tendency for epitope spreading of the autoantibody repertoire.

AA SLE patients had decreased granulocyte, monocyte, pDC, and IL1 signatures and this is likely related to the ancestry associated benign neutropenia, as healthy AA also had these signatures decreased compared to healthy EA. Many AA SLE patients had increased B cells compared to HC, and this suggests some dysregulation in the B cell compartment as there were no increased T cells associated with AA ancestry, and T cells would be just as likely to be affected by the proportional decrease in myeloid cells as a result of benign neutropenia. Whereas all of the increased LDG and monocyte signatures initially detected in NAA turned out to be associated with corticosteroid usage, NAA was positively associated with increased inflammasome, erythrocytes, and the unfolded protein response and negatively associated with IFN, T cells and MHC class II. The NAA association with erythrocytes is of note as an association of SLE and erythrocyte transcripts has been reported but could be related to ancestral background(27). EA was positively associated with low platelets, myeloid secreted, inflammasome, NK cells and the SNOR low down signature, a set of genes overexpressed to SLE patients in the ILL1 and ILL2 clinical trials that initially grouped by the first principal component analysis with HC, but could distinguish this group from HC if compared without the other SLE patients.

Previous work has suggested a strong association between the IGS and autoantibodies(22), and the association of dsDNA with increased plasma cells(46) and T-regs(47) by flow cytometry. Our finding demonstrated that it is not the IGS that is ancestry dependent per se as previously reported(26), but the presence of autoantibodies to RNP/Sm and the increased combination of autoantibodies that is associated with ancestry. Our findings demonstrated that AA patients are likely to have multiple autoantibodies in combination with anti-RNP autoantibodies, and in patients of any ancestry, more autoantibodies and anti-RNP autoantibodies were associated not only with an increased IGS and plasma cell signatures, but also T-reg, cell cycle, and myeloid inflammation signatures. Previous work that did not find an increased association of anti-RNP with the IGS in AA SLE patients(40) is likely related to considering the autoantibodies one at a time instead of in combination. In addition to increasing our understanding of AA SLE, this work has strong implications for using anti-dsDNA to balance cohorts for clinical trial enrollment. The AA SLE patients entered into ILL1 and ILL2 looked similar by anti-dsDNA autoantibodies, but our work showed that this served to severely underestimate the contribution to the transcriptome, and potentially to the disease severity of AA SLE patients. Because being single positive (for the five autoantibodies measured) was the most common finding for the 1100 EA SLE patients in the ILL1 and ILL2 phase 3 clinical trials, it suggests that anti-ds DNA is a good metric for EA autoantibodies, but not AA or NAA autoantibodies.

Importantly, this work considered the combinations of the five autoantibodies to determine the effect of multiple autoantibodies on transcriptomic signatures.The significantly increased IGS in SLE patients of all ancestries with multiple autoantibodies and the almost complete lack of the IGS in the 273 SLE patients without anti-RNP, - dsDNA, -Sm, -SSA or -SSB provides support for the hypothesis that the IGS arises from downstream pattern recognition receptor signaling induced by endosomal TLR7, TLR8 and TLR9 binding to single and double stranded RNA and dsDNA containing immune complexes as previously suggested(48). Autoantibody profiles may be heritable and autoantibody associations for AA SLE patients have been demonstrated for alleles of *LRRC20, LPAR1, EFNA5* and *VSIG2* to anti-SSB, anti-SSA/Sm, anti-RNP and anti-RNP/Sm negative, respectively(49). IFN appears to positively regulate TLR7 signaling and negatively regulate TLR9 signaling suggesting that in the case of chronic stimulation, RNA ligands for TLR7 will augment the IGS and dsDNA ligands will dampen the IGS (50, 51). Another potential contribution to the increased IFN signatures in patients with anti-RNP autoantibodies may be the extrusion of interferonogenic, oxidized mitochondrial DNA by neutrophils in response to anti-Sm/RNP autoantibodies(52, 53). Anti-RNP, -SSA, -Sm and -SSB autoantibodies were also found more commonly in circulating immune complexes compared to anti-dsDNA autoantibodies and immune complex endocytosis by Fc receptors may lead to efficient engagement of TLRs in endosomes and downstream IFN production(54).

Despite the impact of SoC drugs and serologic abnormalities, a clear role for ancestry could be discerned. By stepwise logistic regression analysis, AA was independently associated with increased plasma cell, Ig, and B cell transcripts when modeled alongside autoantibodies, and this, combined with the data from healthy AA subjects, suggests that AA SLE patients have higher background levels of these transcripts. Increased B cell counts and platelets previously were demonstrated (55)(56) to be increased in AA compared to EA SLE subjects. B cell hyperactivity has been proposed as a reason that AA SLE patients have more plasma cells and reactivities to autoantigens, but further work to understand the mechanisms underlying the relationship between the increased B cell signatures and autoantibodies is necessary(40). Reticulocytosis, which may account for the erythrocyte gene transcripts detected in our study, may be augmented in AA SLE patients because the ancestral *G6PD* deficiency may lead to induced hemolysis secondary to infection and leukocyte phagocytosis(57). AA was also associated with decreased granulocyte, monocyte, pDC, anti-inflammation and IL1 signatures and this is likely related to the well described Duffy Null Polymorphism (*ACRK1*) in AA(36, 58). Whereas the increased LDG, granulocyte and monocyte signatures initially detected in NAA turned out to be associated with corticosteroid usage, NAA was positively associated with increased inflammasome and erythrocyte, and with low T cell and MHC class II signatures. The NAA and AA association with erythrocyte transcripts is of note as an association of SLE and erythrocyte transcripts has been reported but could be related to ancestral background(27). NAA SLE patients in this study were receiving more corticosteroids and IS therapy than EA and AA SLE patients and the potent effect of SoC drugs was noted in the high number of discrepant DEGs between NAA and AA or EA. EA was positively associated with low platelets, myeloid secreted, inflammasome, NK cells and the SNOR low down signature. Several published ancestral related genes divergent between AA and EA that are also involved in immune responses were differentially expressed between HC of different ancestries, including *IL8, CXCL1, CXCL5, STAT1, CEPBP, ITGAM* and CD58,(15) providing evidence that ancestral SNPs contribute to the gene expression profile.

This study highlights the importance of appropriate controls for gene expression studies, as the ancestral transcriptomic backbone may be emphasized depending on HC comparators. DEGs might be inappropriately attributed to the disease instead of the ancestry whether or not the allelic differences play an actual role in the pathogenesis of SLE. The ancestral differences between males also appeared similar to the ancestral differences between females suggesting the ancestral component to gene expression will be much more important to take into consideration than male/female differences. Major differences were reported in one lupus cohort between male and female SLE patients with respect to renal involvement and serological manifestations(59), but we detected few gene expression differences between males and females of EA ancestry when matched for SoC drugs.

SoC therapies affected every gene expression signature and accounting for these effects is necessary to interpret blood transcriptomic signatures. SoC drug effects on the transcriptome were confirmed by reports in the literature for the elimination of circulating plasma cells by MTX and MMF (60, 61), elimination of NK cells by AZA(62), and an increase in circulating neutrophils by corticosteroids (63). In what may seem to contrast with previous reports(64, 65) we detected no association between the IGS and anti-malarials; however, previous work looked at IFN protein and not the downstream signature which may be retained in monocytes after the removal of IFN(33). NSAIDs have also been shown to block caspases and inflammation(66) and although the change in GSVA score was not greater than .2, there did appear to be a significant decrease in LDGs and the anti-inflammation signature, at least in EA SLE patients. Corticosteroid usage had a significant effect on most myeloid-related gene signatures and the most potent effect was on the LDG signature with an OR of 3.8. This finding is in contrast to the proposed inflammatory role of LDGs in autoimmunity obtained from in vitro experiments(39, 52, 67). The relationship of corticosteroids to LDGs has strong implications against using this signature as a measure of disease severity or in interpreting LDGs as playing a role in worsening disease as worsening disease might prompt an increase in corticosteroid doses.

It is important to emphasize that common signatures specific for SLE were detected and included genes associated with plasma cell, Ig, IGS, anti-inflammation, cell cycle, T-reg cell, dendritic cell, TNF and myeloid secreted signatures. The balance of these SLE-related abnormalities was different in the various ancestral groups and their prominence was clearly influenced by SoC medications. Despite this, when these influences were considered and mitigated, a set of molecular abnormalities consistent with SLE was discerned, as has been previously suggested(27, 33, 68). However, the interpretation of perturbations in gene expression profiles in subjects with SLE requires that all the individual influences, including ancestry, drug therapy and serological manifestations be considered, as each can have complex and often contradictory effects. Results from single cell technology will also be affected by ancestry and SoC therapy and it will be important to separate out cell populations prominent in ancestries and induced or repressed by concomitant drugs, from cells actively participating in disease processes. Deconvolution of transcriptome data using ancestral, SoC drug, serologic impact and SLE specific signatures has the potential to stratify patients more effectively for therapy or entrance into clinical trials.

## Methods

### SLE Patients

Two large phase 3 clinical trial databases with baseline microarray analysis were analyzed (GSE88884(31)). The ILL1 and ILL2 clinical trials had microarray expression data for 1566 female patients of self-described ancestry: AA (n = 216), EA (n = 1118) and NAA (mostly from South America (n = 232); top three countries of origin Peru (n = 81), Ecuador (n = 30), and Guatemala (n = 27)) and 124 male patients of self-described ancestry: AA (14), EA (93), NAA (17). Patients of other ancestries were removed to avoid low numbers of patients. Ancestral backgrounds were split evenly between the ILL1 and ILL2 datasets, allowing for a training and test set to determine gene expression differences. All patients had a positive anti-nuclear autoantibody (ANA) test, similar disease activity and percentage of patients with anti-dsDNA(30, 32) (**Supplemental Table 1**). The trials excluded patients with progressive lupus nephritis. Most patients recruited had a mixture of six SLE manifestations: arthritis (86.4%), anti-dsDNA (57.5%), low C (40.0%), alopecia (58.9%), rash (68.3%), and mucosal ulcers (31.7%) (**Supplemental Table 2**). The clinical trial database was made available by M.D. Linnick from Eli Lilly. SLE dataset GSE45291 was also analyzed as two cohorts separated by SLEDAI. The first cohort was 73 AA and 71 EA SLE patients with the same range of SLEDAI scores (2 – 11), similar mean SLEDAI (AA 3.78 +/- 2.46; EA 3.53 +/- 2.08) and mode of SLEDAI (2) (**Supplemental Table 3**). The second cohort were 25 AA and 75 EA, all with SLEDAI values of zero. M.A. Petri provided clinical and SoC drug information for dataset GSE45291.

### Gene Expression Datasets

Data were derived from publicly available datasets on Gene Expression Omnibus (GEO, https://www.ncbi.nlm.nih.gov/geo/). Raw data sources are as follows: GSE88884 female whole blood ILL1 (10 female HC, 798 SLE (540 EA, 101 AA, 157 NAA); all with SLEDAI ≥ 6), GSE88884 female whole blood Illuminate 2 (ILL2; 7 female HC, 768 female SLE (578 EA, 115 AA, 75 NAA) all with SLEDAI ≥ 6), GSE88884 male whole blood ILL1 SLE (5 male HC, 59 male SLE (6 AA, 42 EA, 11 NAA), GSE88884 male whole blood ILL2 (4 male HC, 65 male SLE (8 AA, 51 EA, 6 NAA); (GSE45291 whole blood (9 female HC, female SLE: 73 AA, 71 EA with SLEDAI 2 - 11), GSE45291 whole blood (9 female HC, female SLE: 25 AA, 75 EA; all with SLEDAI equal to zero), GSE35846 whole blood from healthy females (55 EA, 22 AA), and GSE111368 whole blood from healthy females (10 AA, 57 EA).

### Quality Control and Normalization of Raw Data Files

Statistical analysis was conducted using R and relevant Bioconductor packages. For datasets GSE88884 (Affymetrix Human Transcriptome Array 2.0) and GSE45291 (Affymetrix HT HG-U133+ PM), non-normalized arrays were inspected for visual artifacts or poor RNA hybridization using Affy QC plots. To increase the probability of identifying differentially expressed genes (DEGs), analysis was conducted using normalized datasets prepared using both the native Affy chip definition files, followed by custom Brain Array Entrez CDFs maintained by the University of Michigan Molecular and Behavioral Neuroscience Institute. The Affy CDFs include multiple probes per gene and almost twice as many probes as BA CDFs. Whereas Affy chip definition files can provide the greatest amount of variance information for Bayesian fitting, the Brain Array chip definition files are used to exclude probes with known non-specific binding and those shown by quarterly BLASTs to no longer fall within the target gene. Illumina CDFs were used for the two Illumina HumanHT-12 V4.0 datasets (GSE35846, GSE111368).

### Differential Gene Expression (DE)

GCRMA normalized expression values were variance corrected using local empirical Bayesian shrinkage before calculation of DE using the ebayes function in the open source BioConductor LIMMA package(69) (https://www.bioconductor.org/packages/release/bioc/html/limma.html). Resulting p-values were adjusted for multiple hypothesis testing and filtered to retain DE probes with an FDR < 0.05(70).

### Determination of Female and Male Patients and Controls

Log2 expression values were used to determine sex of unknown healthy controls and to compute sex module scores using the formula sex module = *XIST*log2expression + *TSIX*log2expression – (*UTY*log2expression + *RPS4Y1* log2expression + *USP9Y*log2expression). Female controls scored above zero and male controls scored below zero. Five SLE patients (three male and two female) with reported sex in GSE88884 ILL1 clinical trial database were found to have expression of genes consistent with the opposite sex (**Supplemental Figure 7**). For all analyses shown in this paper, patients were analyzed with their reported sex in the Illuminate clinical trial database. For **Supplemental Figure 4** analysis of the gene expression differences between males and females, three males with inconsistent sex chromosome gene expression were removed and did not change the results.

### Gene Set Variation Analysis (GSVA)

GSVA(35) (V1.25.0) is an open source software package available from R/Bioconductor and was used as a non-parametric, unsupervised method for estimating the variation of pre-defined gene sets in samples of microarray expression data sets (www.bioconductor.org/packages/release/bioc/html/GSVA.html). The inputs for the GSVA algorithm were a gene expression matrix of log2 microarray expression values for pre-defined gene sets co-expressed in SLE datasets (**Supplemental Table 5**). Enrichment scores (GSVA scores) were calculated non-parametrically using a Kolmogorov Smirnoff (KS)-like random walk statistic and a negative value for a particular sample and gene set, meaning that the gene set has a lower expression than the same gene set with a positive value. The enrichment scores were the largest positive and negative random walk deviations from zero, respectively, for a specific sample and gene set. The positive and negative enrichment score for a particular gene set depend on the expression levels of the genes that form the pre-defined gene set. GSVA calculates enrichment scores using the log2 expression values for a group of genes in each SLE patient and healthy control and normalizes these scores between -1 (no enrichment) and +1 (enriched).

Enrichment modules containing cell type and process specific genes were created through an iterative process of identifying DE transcripts pertaining to a restricted profile of hematopoietic cells in 13 SLE microarray datasets and checked for expression in purified T cells, B cells and monocytes to remove transcripts indicative of multiple cell types as previously described. Genes were identified through literature mining, GO biological pathways, and STRING interactome analysis as belonging to specific categories(33). The TCRA, TCRB, TCRAJ, TCRD, TCRG, and Ig gene lists were taken from the Affymetrix HTA2.0 chip definition. SNOR down low were the seven most decreased transcripts and SNOR up low were the seven most increased transcripts compared to HC for 348 female patients from ILL1 and ILL2 SLE patients that did not separate from HC by principal component analysis (PCA) (FDR < .05) The LDG signature was taken from purified LDGs DE to HC and SLE neutrophils(67) and consists mainly of neutrophil granule proteins from Module B as described in Kegerreis et al(43). The overlap in genes between some signatures was intentional and used to check that signatures were behaving cohesively in patients.

### Weighted Gene Co-expression Network Analysis

Weighted Gene Co-expression Network Analysis (WGCNA)(71) is an open source R package https://horvath.genetics.ucla.edu/html/CoexpressionNetwork/Rpackages/WGCNA/. Log2 normalized microarray expression values for the GSE88884 dataset ILL1 and ILL2 cohorts were filtered using an IQR to remove saturated probes with low variability between samples and used as inputs to WGCNA (V1.51). Adjacency co-expression matrices for all probes in a given set were calculated by Pearson’s correlation using signed network type specific formulae. Blockwise network construction was performed using soft threshold power values that were manually selected and specific to each dataset in order to preserve maximal scale free topology of the networks. Resultant dendrograms of correlation networks were trimmed to isolate individual modular groups of probes, labeled using semi-random color assignments, based on a detection cut height of 1, with a merging cut height of 0.2, with the additional use of a partitioning around medoids function. Final membership of probes representing the same gene into modules was based on selection of greatest scale within module correlation against module eigengene (ME) values. Correlation to ancestry was performed using Pearson’s r against MEs, defining modules as either positively or negatively correlated with those traits as a whole.

### Gene Overlap

Gene Overlap is an open source R bioconductor package (www.bioconductor.org/packages/release/bioc/html/GeneOverlap.html) used to test the significance of overlap between two sets of gene lists. It uses the Fisher’s exact test to compute both an OR and overlap p value. For comparison of datasets on different array platforms (Illuminate versus Affymetrix), an FDR < 0.2 was used.

### Stepwise Logistic Regression Modeling

SAS 9.4 (Cary, NC) was used for stepwise logistic regression. GSVA enrichment scores greater or less than healthy control averages plus or minus one standard deviation were determined and SLE patients were assigned a 1 or 0 based on having a signature greater or less (Low) than HC. These scores were used as 34 dependent binary variables to be modeled individually as the outcome variable to 26 independent binary variables: ancestry (AA, EA, NAA), drugs (corticosteroids, anti-malarials, NSAIDs, AZA, MTX, MMF, Cyclophosphamide), SLE manifestations (rash, arthritis, mucosal ulcers, vasculitis, alopecia) autoantibodies and complement (anti-RNP+dsDNA+ plus any of SSA, SSB or Sm, anti-RNP+dsDNA-plus any of SSA, SSB or Sm, anti-RNP-dsDNA+ plus any of SSA, SSB, or Sm, and SSA, SSB or Sm, low C3, low C4) and time (Age > 50, time from onset of disease (≤1 year, >1 year ≤ 5 years, > 5 years ≤ 10 years, > 10 years). Spearman correlation coefficients were determined between variables before stepwise logistic regression in order to determine whether groups were too similar to give independent information to the model (co-linearity). The ethnic term Hispanic as a general category was removed since it had an r_s_ > .5 compared to NAA (**Supplemental Table 17**). The stepwise approach was used to produce the statistically significant model. The results of any model that violated the Hosmer Lemeshow test were discarded. The p values, OR, and CI are listed in **Supplemental Tables 18 - 21**.

### CIRCOS

CIRCOS (V0.69.3) software was used to visualize the OR determined by stepwise logistic regression analysis. OR do not go below zero and a change from an OR of 0.5 to .25 is the same relative change as that between 2.0 and 4.0. For representative visualization, OR between 0 and 1 were converted to the 1/X value where X is an OR between 0 and 1. An interval graph was used to assign thickness of the lines where OR < 2 = 1pt; 2 ≥ OR < 3 = 5pt; 3 ≥ OR < 10 = 10pt; OR ≥ 10 = 20pt.

### Machine Learning Analysis

Logistic regression and two distinct machine learning algorithms to predict the AA SLE patients from EA SLE patients were employed. Logistic regression, an elastic generalized linear model (GLM) and Support Vector Machine (SVM) were deployed to predict the ancestry status of SLE samples and determine the top 25 predictors using the gene importance score. R was used for implementation, as it is an open source statistical language with access to machine learning algorithms. Logistic regression, GLM, and SVM were implemented using glmnet, nnet, e1071 R packages, respectively. The performance of the machine learning models was evaluated by 10-fold cross validation technique. This methodology avoids the problem of over-fitting by using all the observations for both training and validation by randomly assigning each patient to one of 10 groups which results in a less biased model. The model was fit using the first 9 folds for training and validated using the remaining 10th fold for testing. Similarly, each fold was validated. Performance metrics such as sensitivity and specificity were determined by averaging class probabilities from each fold. Receiver Operating Characteristic (ROC) curves and area under curve were plotted and measured for each machine learning model using R.

### Statistics

GraphPad PRISM 8 version 8.2.1 was used to perform mean, median, mode, standard deviation, ANOVA, Tukey’s multiple comparisons test, Sedak’s multiple comparisons test, linear regression analysis and unpaired t-test with Welch’s correction. The Fisher’s exact test was performed in R.

### Data Availability

All microarray datasets in this publication are available on the NCBI’s database Gene Expression Omnibus (GEO) (https://www.ncbi.nlm.nih.gov/geo/).

### Code Availability

All bioinformatic software used in this publication is open source, freely available for R. Additionally, example code used in this paper for LIMMA, GSVA and WGCNA are available at figshare, www.figshare.com. File names are “AMPEL BioSolutions LIMMA Differential Expression Analysis Code”, “AMPEL BioSolutions Gene Set Variation Analysis Code”, and “AMPEL BioSolutions Weighted Correlation Network Analysis WGCNA Code”.

## Data Availability

All microarray datasets in this publication are available on the NCBI database Gene Expression Omnibus (GEO) (https://www.ncbi.nlm.nih.gov/geo/). All bioinformatic software used in this publication is open source, freely available for R. Additionally, code used in this paper for LIMMA, GSVA and WGCNA are available at figshare, www.figshare.com. File names are AMPEL BioSolutions LIMMA Differential Expression Analysis Code, AMPEL BioSolutions Gene Set Variation Analysis Code, and AMPEL BioSolutions Weighted Correlation Network Analysis WGCNA Code.

https://www.ncbi.nlm.nih.gov/gds/?term=GSE88884[Accession]

https://www.ncbi.nlm.nih.gov/geo/query/acc.cgi?acc=GSE45291

https://www.ncbi.nlm.nih.gov/gds/?term=GSE35846

https://www.ncbi.nlm.nih.gov/gds/?term=GSE111368

## Author Contributions

M.D.C. performed data analysis, generated figures, and wrote the manuscript. P.B. and N.S.G. performed data analysis and generated figures. A.C.G. and M.A.P. supervised data analysis. A.E.Y. carried out statistical analysis. M.A.P. provided clinical data for GSE45291 and reviewed the manuscript. P.E.L. supervised data analysis, directed the study and wrote the manuscript.

## Acknowledgments

We thank M.D. Linnik at Lilly for the clinical information for dataset GSE88884.

